# Episode-specific and common intrinsic functional network patterns in bipolar

**DOI:** 10.1101/2024.07.26.24310655

**Authors:** Xiaobo Liu, Zhen-Qi Liu, Bin Wan, Xihan Zhang, Lang Liu, Jinming Xiao, Yao Meng, Xiaoqiang Liu, Sanwang Wang, Chao Weng, Yujun Gao

## Abstract

Understanding the alterations in brain function across different episodes of bipolar disorder (BD), including manic (BipM), depressive (BipD), and remission states (rBD), poses a significant challenge. In our cross-sectional study, we collected resting-state functional magnetic resonance imaging data from 117 BD patients (BipM: 38, BipD: 42, rBD: 37) and 35 healthy controls. Our aim was to delineate functional connections associated with episode dynamics, delineate common and specific patterns, validate them as biomarkers, and elucidate their biological underpinnings. Initially, we identified a common altered pattern within the subregions of the ventral-attention network, alongside specific patterns observed in the default mode network for BipM, the prefrontal network for BipD, and the limbic network for rBD. Using large-sample data from the Human Connectome Project, we further identified that these connectivity patterns exhibit relatively high reliability and heritability. Also, these distinct patterns accurately characterized the diverse episodes of BD and effectively predicted the corresponding clinical symptoms linked with each episode type. Importantly, using out of sample data to decode possible neurobiological mechanisms underlying these patterns, we found that regions of particular interest were enriched in multiple receptors, including MOR, NMDA, and H3 for specific alterations, and A4B2, 5HTT, and 5HT1a for common alterations. Moreover, both episode-specific and common patterns demonstrated a high enrichment for cell types such as L5ET, Micro/PVM,oligodendrites and Chandelier. Our study offers novel insights concerning episode dynamics in BD, paving the way for personalized medicine approaches tailored to address the various episodes.

## INTRODUCTION

Bipolar disorder (BD) is a psychiatric condition characterized by a spectrum of episodes, including manic episodes (BipM), depressive episodes (BipD),remitted bipolar disorder (rBD). BD affects more than 2% of the global population and is associated with high levels of disability, suicide rates, and prevalence (Grande et al. 2016). Patients with BD suffer from various functional impairments across different episode states. For instance, cognitive impairments are more closely linked with BipM rather than BipD or general mood fluctuations (Su et al. 2021). During therapy, individuals with BD experience emotional shifts across various episode states (Pomarol-Clotet et al. 2015). Different stages of episodes have strong commonalities and unique characteristics, understanding how brain functions dynamically change with episodes is crucial for developing effective treatment strategies for BD.

Functional magnetic resonance imaging (fMRI) is a widely used non-invasive tool for studying brain function in BD research (Strakowski et al. 2012, Thermenos et al. 2010). Previous resting-state fMRI studies have identified significant differences between BD patients and control groups in the default mode network (DMN), central executive network (CEN), and emotion regulation networks (Vargas et al. 2013, Wu et al. 2023). Notably, CEN dysfunction has been linked to cognitive impairments in BD patients (Thermenos et al. 2010). Wang et al. (Wang et al. 2020) investigated different episodes of bipolar disorder, reporting decreased connectivity within the affective network during acute episodes, which was not observed during remission. Reduced connectivity within the DMN during acute states was replaced by increased connectivity during remission. These findings reveal issues related to self-reflection and emotion regulation, which manifest differently during manic and depressive episodes (Yoon et al. 2021). Furthermore, a widely accepted underlying mechanism of bipolar disorder involves the regulation of emotional behavior (e.g., white matter connectivity and prefrontal pruning), leading to reduced connectivity between the ventral prefrontal networks and limbic regions (Strakowski et al. 2012). This research suggests that common functional network differences during different episodes may result from shared structural lesions. While these studies provide crucial insights into brain functional changes associated with different episodes of bipolar disorder, a comprehensive comparison among BipM, BipD, rBD, and control groups is needed to fully understand the common and specific alterations. Such understanding could advance more precise treatments for different stages of the disorder.

Neurobiological factors, such as gene expression and neurotransmitter systems related to episode dynamics, have garnered increasing attention. During manic episodes, dopamine levels may rise, while depressive periods might be associated with a decrease in dopamine levels(Ashok et al. 2017). Variations in neurotransmitter levels can lead to diverse activation patterns within brain networks. For instance, altered dopamine signaling can significantly impact functional connectivity in brain regions associated with mood regulation and cognitive function(Robison and Nestler 2011). Heritability and reliability in the functional connectome vary across regions and components, underscoring the genetic influence on brain network organization(Teeuw et al. 2021). Specific gene expression profiles have been linked to the regulation of synaptic plasticity and neural excitability, which are crucial for maintaining stable mood states(Gordovez and McMahon 2020). Despite these insights, the episode-specific and common molecular mechanisms of bipolar disorder remain unknown. Understanding these differences and connections will not only elucidate the potential biological underpinnings of the bipolar spectrum, such as more refined connectivity patterns, but also guide subsequent treatments, molecularly targeted therapies, and probe design.

In this study, we aim to investigate the common and specific network patterns related to episode dynamics in the bipolar spectrum and associate these patterns with spatial patterns of gene expression, brain cellular maps, neurotransmitters, and receptors. **Figure 1** shows an overview of the analytic steps. We first construct resting-state intrinsic functional connectomes in individuals across three bipolar episode states and healthy controls, then categorize the significant connectivity edges into episode-specific and common network patterns. Finally, we filter the patterns using reliability and heritability metrics from a normative sample (Human Connectome Project). To clarify the potential neurobiological processes underlying altered brain functions, we calculated the regional centrality from the common and specific patterns and linked it to the spatial patterns of gene expression, brain cellular maps, neurotransmitters, and receptors, using *neuromaps* (Markello et al. 2022).

**TABLE I.**
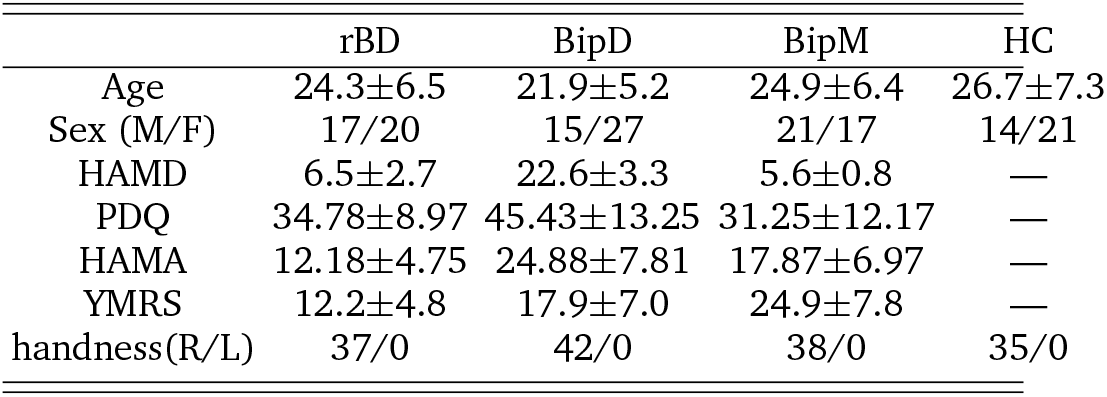
Demography. HAMD means Hamilton Depression Rating Scale, YMRS means Young Mania Rating Scale, PDQ means Perceived Disability Questionnaire, HAMA means Hamilton Anxiety Scale, M means male, F means female, R means right hand, L means left hand.

|  | rBD | BipD | BipM | HC |
| --- | --- | --- | --- | --- |
| Age | 24.3±6.5 | 21.9±5.2 | 24.9±6.4 | 26.7±7.3 |
| Sex (M/F) | 17/20 | 15/27 | 21/17 | 14/21 |
| HAMD | 6.5±2.7 | 22.6±3.3 | 5.6±0.8 | — |
| PDQ | 34.78±8.97 | 45.43±13.25 | 31.25±12.17 | — |
| HAMA | 12.18±4.75 | 24.88±7.81 | 17.87±6.97 | — |
| YMRS | 12.2±4.8 | 17.9±7.0 | 24.9±7.8 | — |
| handness(R/L) | 37/0 | 42/0 | 38/0 | 35/0 |

**Figure 1.**
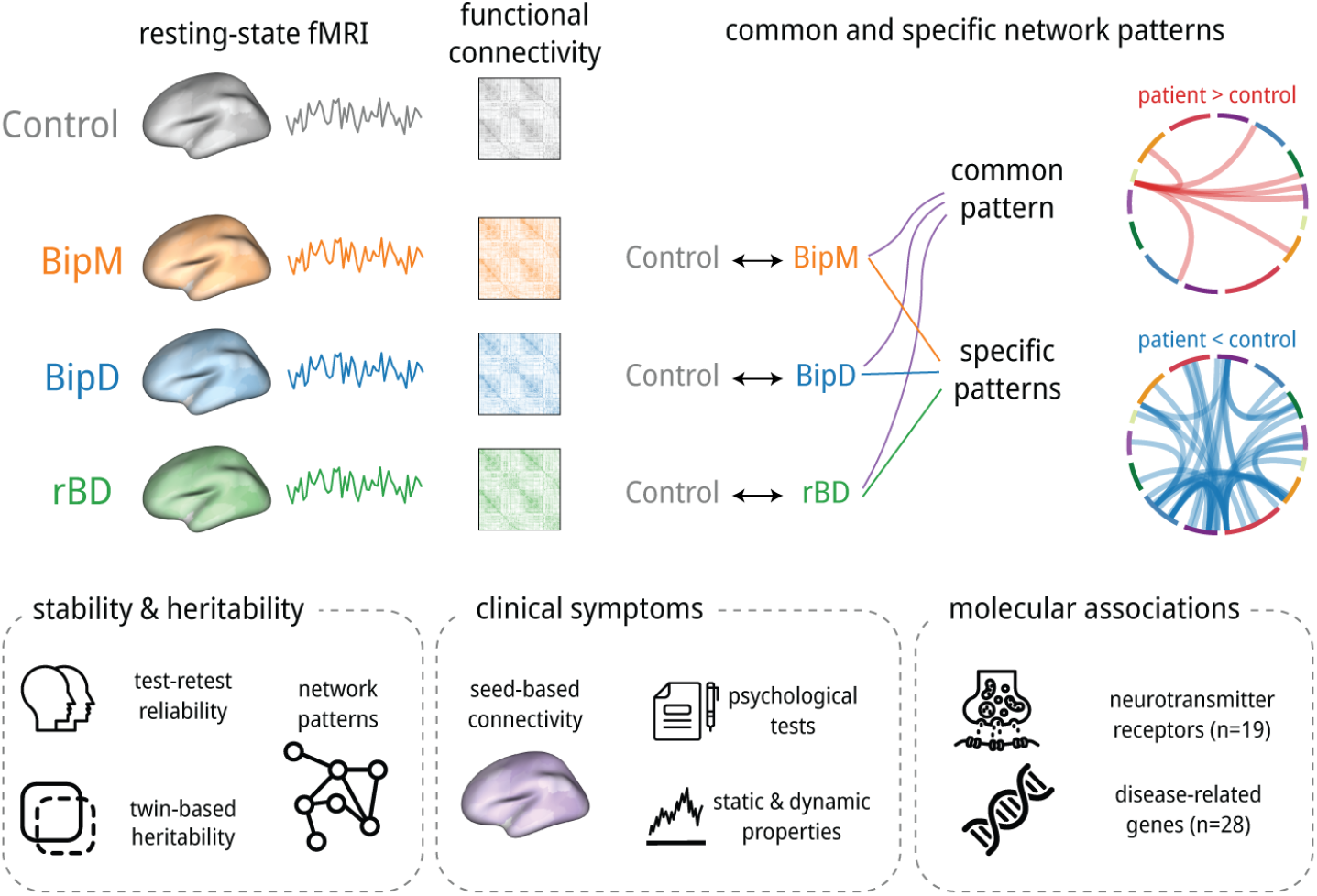
Overview. We first constructed functional connectivity maps and compared them across different disease episode types. Next, we categorized these connectivities into common and specific patterns, which were validated using machine learning techniques. To ensure robustness, we assessed the test-retest reliability and twin-based heritability of these patterns and filtered them accordingly. Subsequently, we linked these network patterns and their properties to clinical symptoms. Finally, we explored the potential molecular mechanisms underlying these patterns.

## RESULTS

### Common and specific network patterns

In our study, we compared the connectivity edges between patients and healthy controls using a two-sample t-test. We subsequently identified significant common and specific patterns by selecting overlapping and non-overlapping edges, respectively. To pinpoint pathological regions, we calculated the degree of significant abnormal networks (**Figure 2**). During the BipM episode, the prefrontal cortex, encompassing the dorsolateral prefrontal cortex (DFC) and medial prefrontal cortex (PFCm) within the default mode network (DMN), along with the parahippocampal cortex (PHC) and orbitofrontal cortex (OFC) within the limbic network, exhibited the most pronounced network differences. In contrast, the BipD episode was characterized by substantial network pattern differences in the posterior cingulate cortex (pCun) of the fronto-cingulate network (FCN), subregions within the visual network (VN), and subregions within the somatomotor networks (SMN). Specific patterns in the rBD group primarily involved the OFC within the limbic network, the precentral visual area (PrCv) of the dorsal attention network (DAN), and subregions within the VN. Common network patterns across these episode phases were predominantly associated with the OFC within the limbic network, the PrCv within the DAN, subregions within the SMN, and the parietal operculum (ParOper) and frontal operculum and insula (FrOperIns) within the ventral attention network (VAN). In summary, our findings indicate that the specific altered networks were the DFC and PFCm of the DMN during BipM, the pCun in the FCN during BipD, and the OFC in the limbic network during rBD. The common pattern was located in the PrCv and FrOperIns in the VAN.

**Figure 2.**
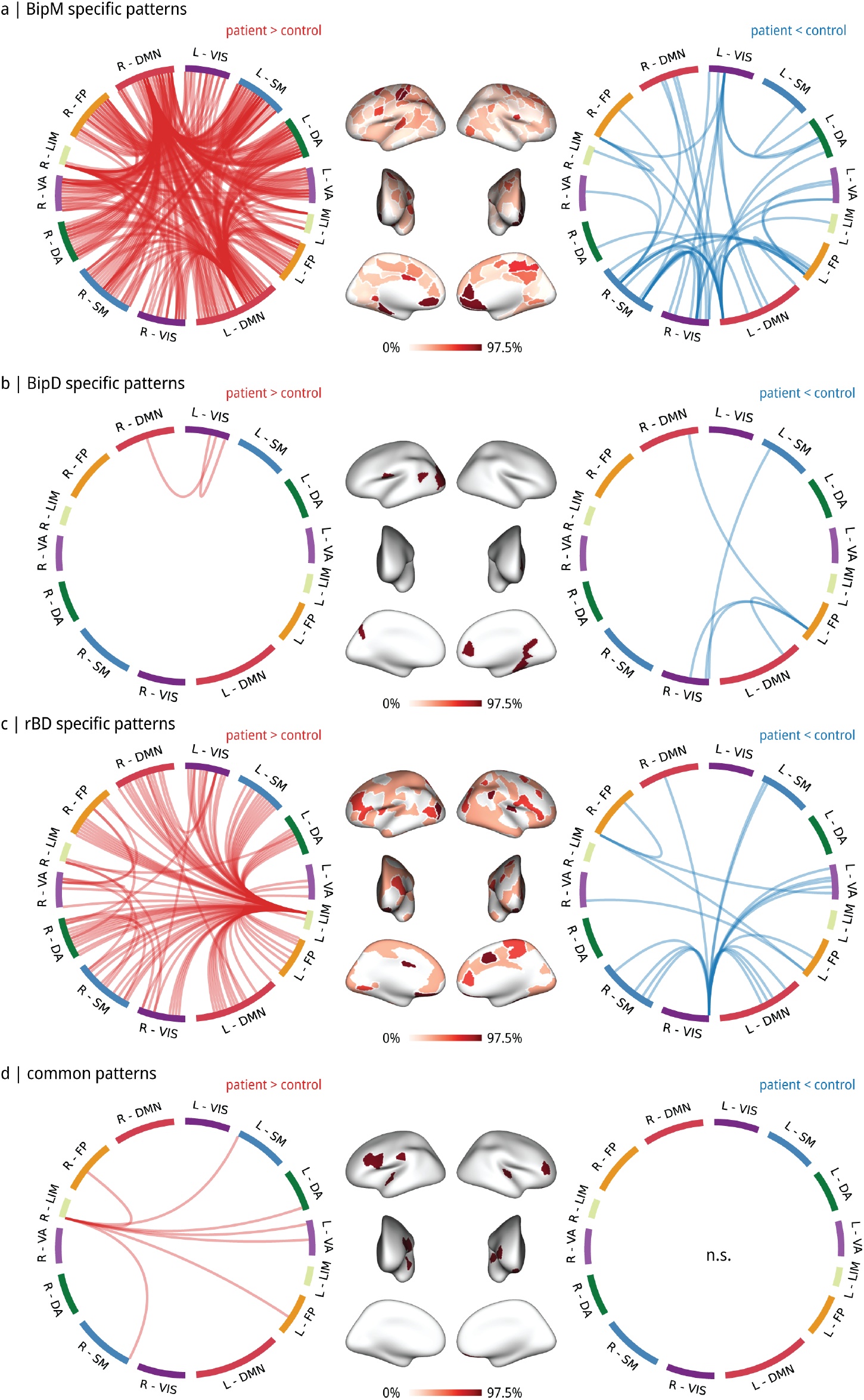
Common and specific network patterns. Specific pathogenic network patterns of BipD **(a)**, BipM **(b)**, and rBD **(c). (d)** common network patterns. During the BipM episode, the greatest network differences were in the prefrontal cortex (DFC and PFCm) of the DMN, and the PHC and OFC of the limbic network. In the BipD episode, the largest differences were in the pCun of the FCN, and subregions in the VN and SMN. Specific patterns in the rBD involved the OFC of the limbic network, the PrCv of the DAN, and subregions in the VN. Common patterns across these phases were predominantly associated with the OFC in the limbic network, the PrCv in the DAN, subregions in the SMN, and the ParOper and FrOperIns in the VAN.

### Filtering patterns

To obtain stable alterations, we filtered the common and specific patterns based on test-retest reliability and heritability in the twin-designed HCP. In particular, we computed the reliability (intraclass correlation coefficient, ICC) and narrow-sense heritability (*h*^2^) for all connectome edges.

We found that 98.07% of specific pattern edges in BipM, 100% in BipD, and 89.15% in rBD were significantly reliable (*P*_FDR_ < 0.05), with average ICCs of 0.48 ± 0.13, 0.50 ± 0.16, and 0.38 ± 0.10, respectively. For the common network pattern, 100% of edges were significantly reliable, with an average ICC of 0.40 ± 0.06.

Regarding heritability, the average heritability of the specific pattern was 0.19 ± 0.091 in rBD, 0.32 ± 0.086 in BipD, and 0.27 ± 0.09 in BipM. The average heritability of the common pattern was 0.13 ± 0.071. Additionally, the proportion of significant edges (*P*_FDR_ < 0.05) in the specific pattern was 88.37% in rBD, 100% in BipD, and 94.93% in BipM. However, only 57.14% of the common pattern showed statistical significance. Therefore, we selected both significantly stable and significantly heritable edges as the filter for network patterns, and nodes with the largest degree remained consistent (**Figure 3**).

**Figure 3.**
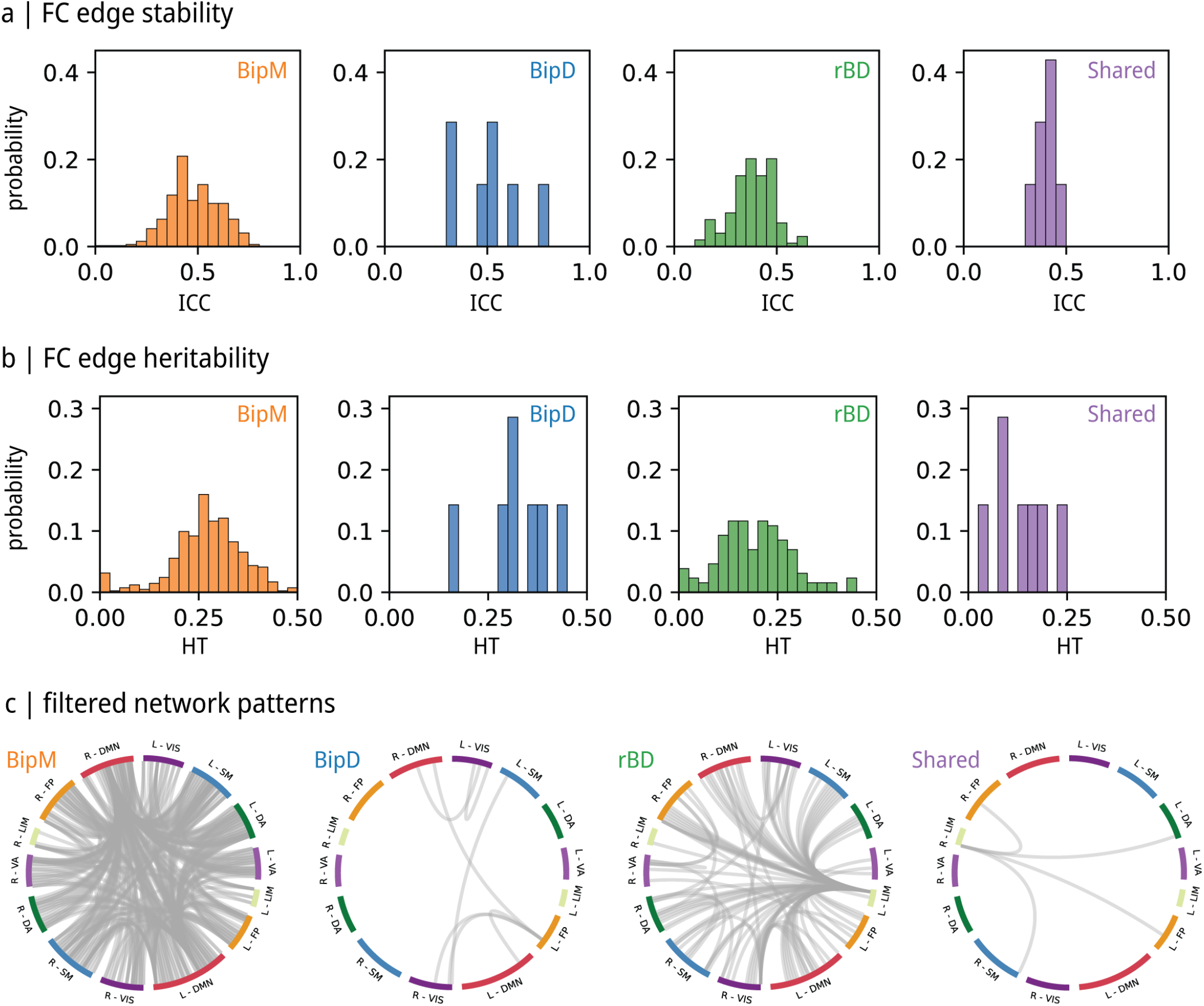
Reliability and heritability of Common and specific patterns via HCP test-retest dataset and twins dataset. **(a)** Reliability of FCs patterns. Note that the histogram represents the proportion of significant edges within specific networks. **(b)** Heritability of FCs patterns. Note that the histogram represents the proportion of significant edges within specific networks.**(c)** filtered network patterns. We selected significantly stable and significantly heritable edges.

**Figure 4.**
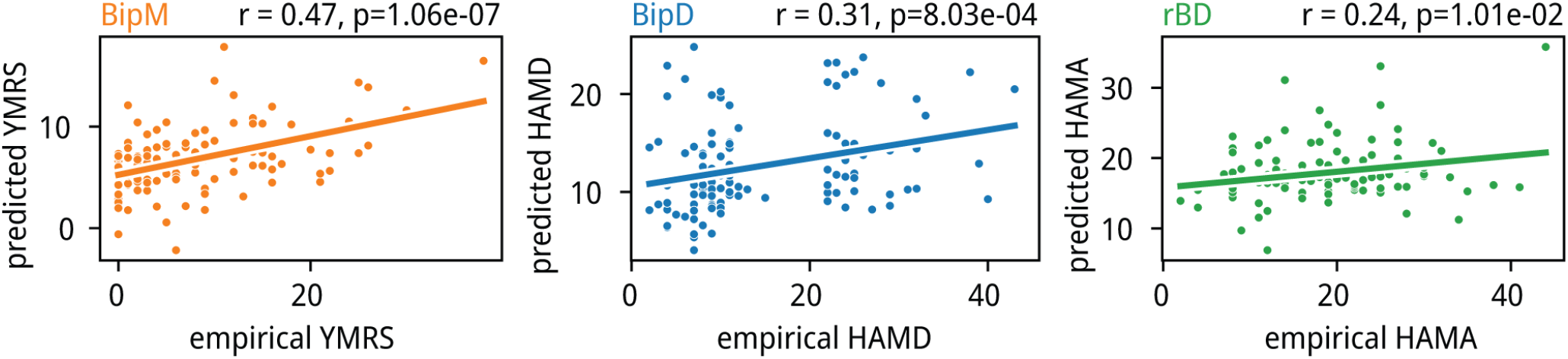
Specific network patterns of various episodes predicted clinical symptom. We implemented the SVR algorithm to predict clinical symptoms via the specific network of Episode phases of bipolar disorder. We found significant correlations between SVR-based predicted score and empirical score (for YMRS: p = 1.06e-07, r = 0.47, for HAMD: p = 8.03e-04, r = 0.31, for HAMA p =1.01e-02, r = 0.24).

**Figure 5.**
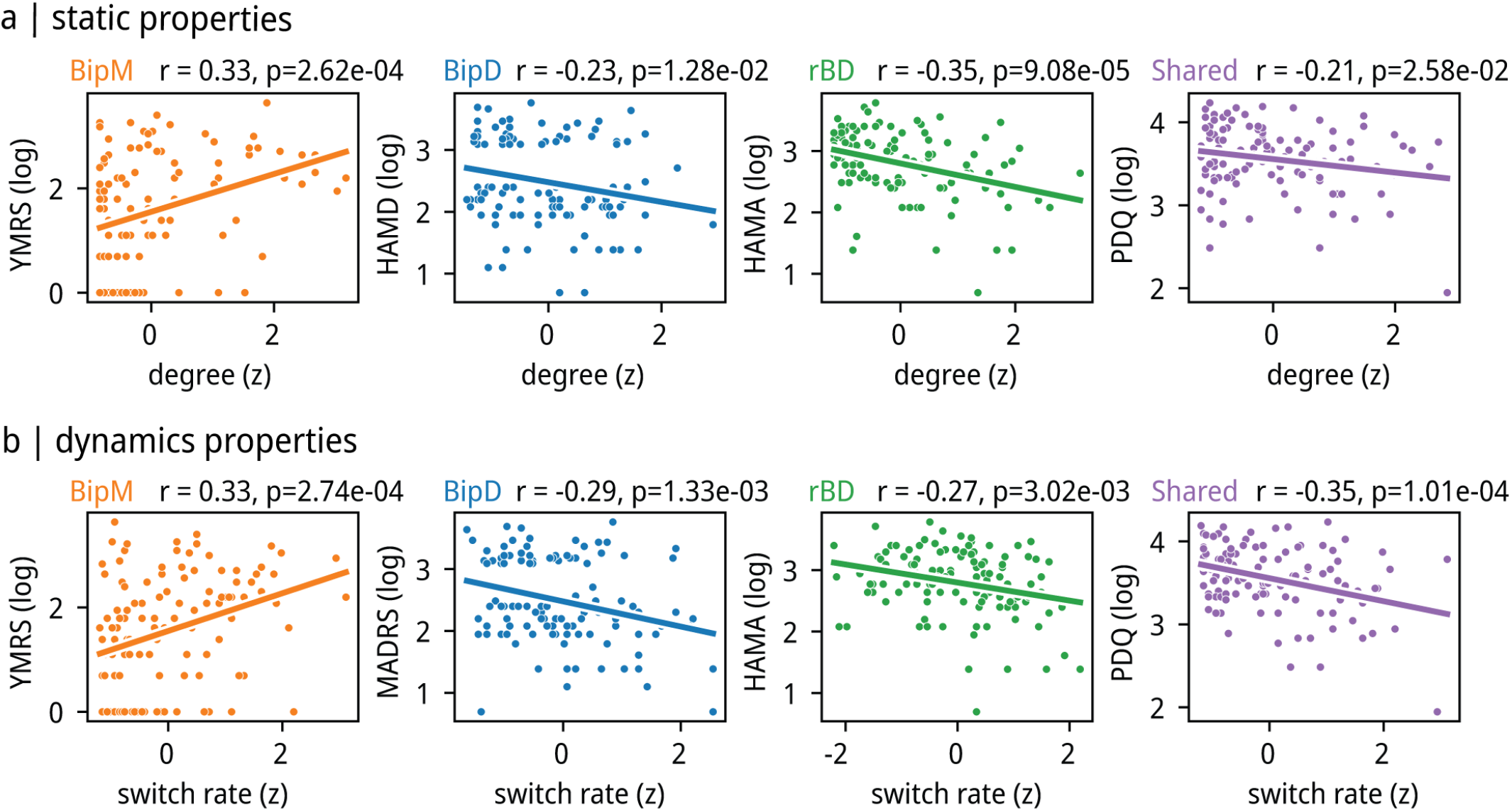
Static and dynamic properties of specific-common hubs. **(a)** The correlation between average degree and cognition in the hubs of both specific network patterns and shared network patterns. Specifically, the observed correlation was significantly negative between degree and HAMA score (p = 0.017, r =- 0.22) in rBD hub, PDQ scores in shared hub (p = 0.019, r = - 0.22) as well as degree and HAMD score and average degree in BipD hub (p = 0.012, r = - 0.23). Also, we found a significantly positive relationship between degree and YMRS score (p = 3.94 × 10-5, r = 0.37) in the BipM hub. **(b)** Average switch rate in the hubs of both specific network patterns and common network patterns and cognition. We found a significant negative relationship between the HAMD score and average switch rate (p = 0.0050, r = -0.26) in the BipD hub, average switch rate in the shared hub (p = 0.0011, r = - 0.30), PDQ score and HAMA score and average switch rate in rBD hubs (p = 0.0077, r = -0.25). Also, we found a positive relationship between the YMRS score and the average switch rate (p = 0.0036, r = 0.27) in the BipM hub.

**Figure 6.**
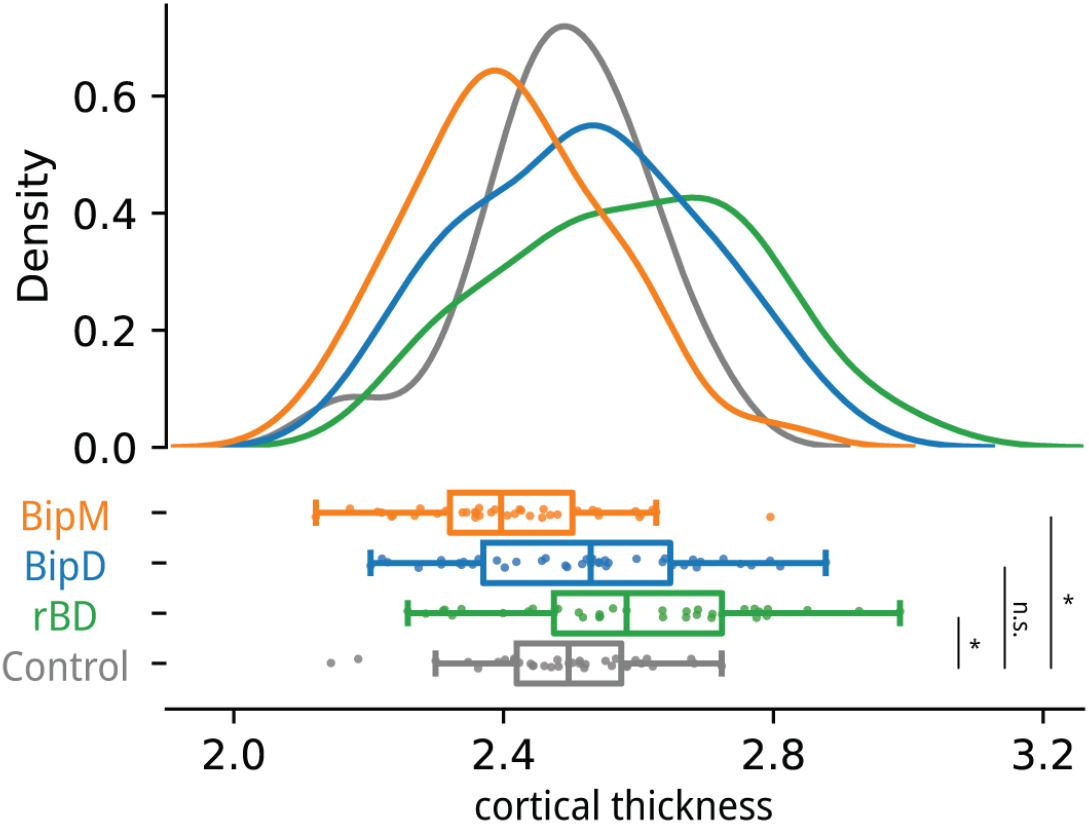
The cortical thickness of rBD, BipD, BipM, and HC in common regions. **(a)** We further investigated common functional patterns with similar structural foundations; thus, we compared the cortical thickness of the common nodes across different episode types of specific diseases. We observed a significantly increased cortical thickness in rBD compared to HC (p = 0.0071, t = 2.77), while BipM exhibited a significantly lower cortical thickness than HC (p = 0.0071, t =-2.77). However, no significant difference in cortical thickness was found between BipD and healthy controls (p = 0.39, t = 0.87). Correspondingly, the cortical thickness of rBD showed no significant difference from BipD (p = 0.071, t = 1.83) but was significantly higher than BipM (p = 0.000011, t = 4.72). Similarly, BipD exhibited a significantly higher cortical thickness than BipM (p = 0.0036, t = 3.01).

**Figure 7.**
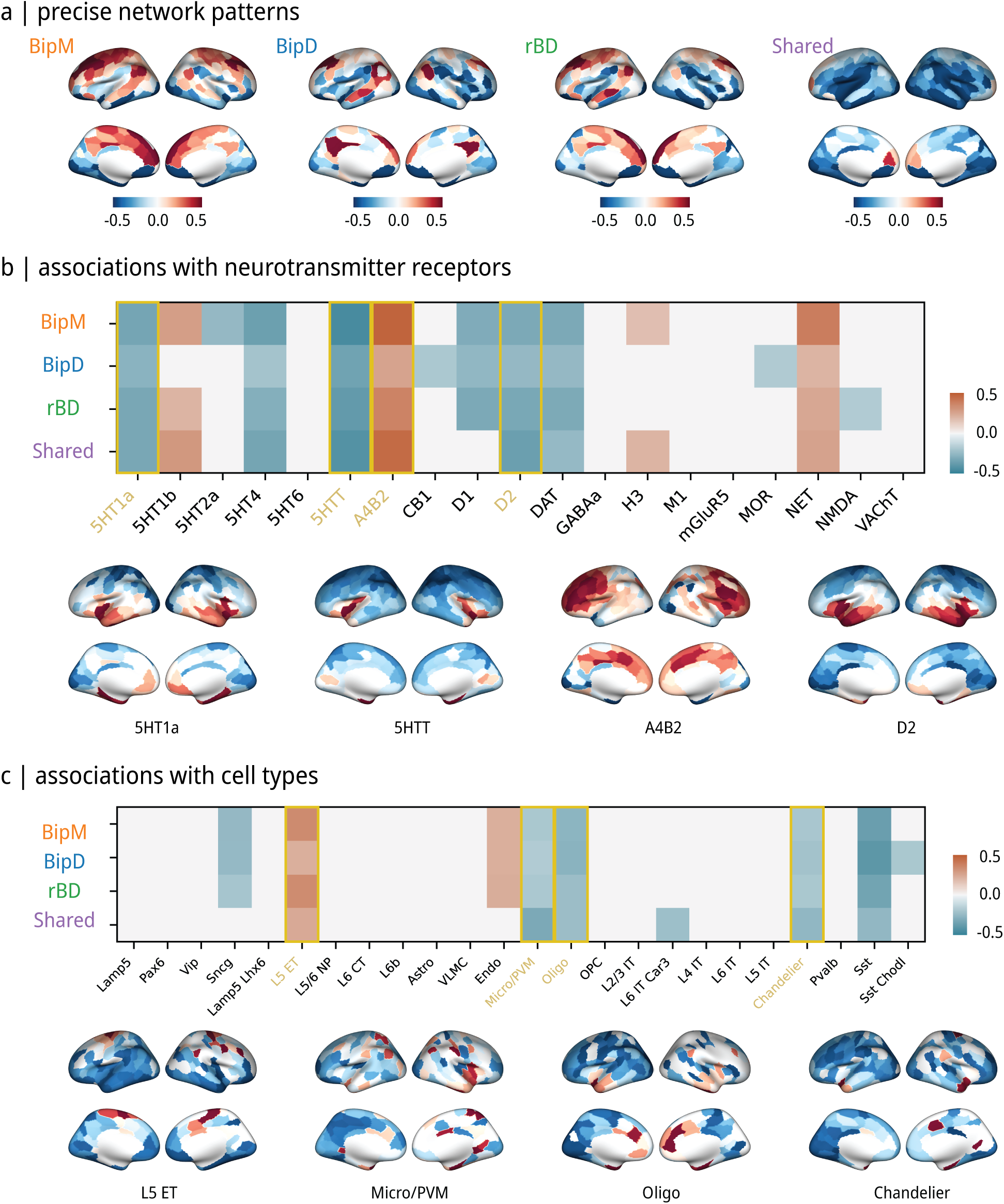
Molecular Structure of specific-Common regions based functional patterns. **(a)** Average seed-based network patterns. **(b)** Receptor mechanism of specific network patterns. During the remission phase, networks were identified that exhibited associations with receptors, including 5HT1a, 5HT1b, 5HT4, 5HTT, A4B2, D1, D2, DAT, NET, and NMDA. In the depressive phase, networks were correlated with receptors such as 5HT1a, 5HT4, 5HTT, A4B2, CB1, D1, D2, DAT, MOR, and NET. Manic phase networks displayed relationships with receptors 5HT1a, 5HT1b, 5HT2a, 5HT4, 5HTT, A4B2, D1, D2, DAT, H3, and NET. Shared networks across phases involved receptors 5HT1a, 5HT1b, 5HT4, 5HTT, A4B2, D2, DAT, H3, and NET. **(c)** Cellular mechanism of specific network patterns. At the cellular level, both common and specific patterns were associated with L5ET, Micro/PVM, Oligo, and Chandelier maps.

### Cross-episodes validation

To validate the network patterns, we implemented cross-episode validation to classify patients and healthy controls using both common and specific patterns (see Table S1 and Table S2). The specific network for BipD showed a classification accuracy of 80.56% (with an AUC of 0.87, *P*_null_ < 0.05). The BipD-specific pattern achieved recognition rates of 59.74% (with an AUC of 0.68) in BipM and 56.16% (with an AUC of 0.61) in rBD. For the specific network of BipM, the classification accuracy was 71.43% (with an AUC of 0.79, *P* null < 0.05), with recognition rates of 51.39% (with an AUC of 0.62) in BipD and 52.05% (with an AUC of 0.64) in rBD. The specific network for rBD had a classification accuracy of 75.34% (with an AUC of 0.80, *P* null < 0.05), with recognition rates of 51.39% (with an AUC of 0.50) in BipD and 58.44% (with an AUC of 0.64) in BipM.The shared patterns were effective in identifying all mood episode types, with accuracies of 75% (with an AUC of 0.84) in BipD, 72.73% (with an AUC of 0.78) in BipM, and 79.45% (with an AUC of 0.86) in rBD.

### Specific network patterns predicted clinical symptoms

We implemented a machine learning algorithm to predict clinical symptoms using the extracted network patterns of specific episodes of BP (see Methods). The results showed a significant correlation between the predicted and empirical YMRS scores using BipM-specific networks (*r* = 0.47, *P* = 1.06e-7), between the predicted and empirical HADRS scores using BipD-specific networks (*r* = 0.31, *P* = 8.03e-4), and between the predicted and real HAMA scores using rBD-specific networks (*r* = 0.24, *P* = 1.01e-2).

### Static and dynamic structure of specific and common network pattern and clinical symptom

In the context of rBD, our investigation revealed noteworthy correlations within specific hubs. Specifically, a statistically significant positive correlation was identified between degree and HAMA score in the rBD hub (*r* = 0.28, *P* = 0.002). Similarly, in the BipM hub, there was a significantly positive correlation between degree and YMRS score (*r* = 0.37, *P* < 0.001), while in the shared hub, a positive correlation was found between degree and PDQ scores (*r* = 0.24, *P* = 0.009). Additionally, within the BipD hub, a significant negative relationship emerged between the HAMD score and average degree (*r* = -0.23, *P* = 0.012).

Moving on to the assessment of average switch rates within both specific network patterns and shared network patterns alongside cognitive factors, intriguing patterns emerged. Notably, a significant negative association was observed between the HAMD score and the average switch rate in the BipD hub (*r* = -0.25, *P* = 0.005). Similarly, in the BipM hub, a negative correlation was found between the YMRS score and average switch rate (*r* = -0.30, *P* = 0.001), and in the shared hub, a negative relationship was identified between the PDQ score and average switch rate (*r* = -0.30, *P* = 0.0011). Additionally, a positive relationship surfaced between the HAMA score and the average switch rate in rBD hubs (*r* = 0.30, *P* = 0.001).

### Structural basis of common networks

We conducted an in-depth investigation into common nodes characterized by similar structural foundations. Subsequently, we undertook a comparative analysis of cortical thickness within the default mode network across diverse episode types of specific diseases. Our findings revealed a notable increase in cortical thickness in individuals with rBD compared to HC (*t*= 2.77, *P* = 0.007). Conversely, BipM manifested a significantly diminished cortical thickness relative to HC (*t* = 2.77, *P* = 0.007). Notably, no statistically significant specification in cortical thickness emerged between individuals with BipD and the HC group (*t* = 0.87, *P* = 0.390). Moreover, the cortical thickness of rBD exhibited no significant disparity compared to BipD (*t* = 1.83, *P* = 0.071) but demonstrated a significant elevation compared to BipM (*t* = 4.72, *P* < 0.001). Likewise, BipD exhibited a significantly higher cortical thickness compared to BipM (*t* = 3.01, *P* = 0.004).

### Molecular mechanisms of specific and common network patterns

Finally, we conducted a thorough analysis of the molecular and genetic foundations associated with specific and shared patterns during different phases of bipolar disorder. Notably, during the remission phase, receptors including 5HT1a, 5HT1b, 5HT4, 5HTT, A4B2, D1, D2, DAT, NET, and NMDA were found to be significantly linked to specific networks (*P*_FDR_ < 0.05, *P*_*spin*_ < 0.05). Similarly, in the depressive phase, specific networks (*P*_FDR_ < 0.05, *P*_spin_ < 0.05) were associated with receptors such as 5HT1a, 5HT4, 5HTT, A4B2, CB1, D1, D2, DAT, MOR, and NET. Networks specific to the manic phase (*P*_FDR_ < 0.05, *P*_spin_ < 0.05) were related to receptors, including 5HT1a, 5HT1b, 5HT2a, 5HT4, 5HTT, A4B2, D1, D2, DAT, H3, and NET. Furthermore, receptors associated with shared network patterns (*P*_FDR_ < 0.05, *P*_spin_ < 0.05) across phases included 5HT1a, 5HT1b, 5HT4, 5HTT, A4B2, D2, DAT, H3, and NET. Additionally, our findings revealed that 5HT1a, 5HT4, 5HTT, A4B2, D1, D2, DAT, and NET are implicated in both specific and common networks across different phases of bipolar disorder. NMDA was significantly related to rBD, MOR was significantly related to BipD, and H3 was significantly related to BipM. At the cellular level, we found both common and specific patterns were significantly associated with the spatial distribution of layer 5 extratelencephalic-projecting excitatory neuron (L5ET), microglia/Perivascular macrophage (Micro/PVM), oligodendrocyte (Oligo), and Chandelier parvalbumin-expressing interneuron (*P*_FDR_ < 0.05, *P*_spin_ < 0.05) Jorstad et al. (2023), Zhang et al. (2023).

On the genetic level, during the remission phase, specific network patterns were associated (*P*_FDR_ < 0.05, *P*_spin_ < 0.05) with PLEKHO1, SCN2A, POU3F2, and ANK3. Conversely, the manic phase (*P*_FDR_ < 0.05, *P*_spin_ < 0.05) exhibited specific network patterns associated with SCN2A, POU3F2, and ANK3. Common network patterns were identified with PLEKHO1, LMAN2L, SCN2A, and POU3F2 (*P*_FDR_ < 0.05, *P*_spin_ < 0.05). Notably, no specific network patterns related to the depressive phase were found to be associated with genes implicated in bipolar disorder.

## DISCUSSION

In the present study, we uncovered dynamic intrinsic network alterations in BD. A common altered pattern consistent across three episodes was primarily observed in the VAN. Specifically, BipM exhibited alterations predominantly within the DMN, BipD displayed aberrations mainly in regions of the FCN, and rBD showed disruptions in subregions of the limbic network. Machine learning classification suggested that specific patterns contribute to the distinct episodes. Furthermore, by analyzing test-retest reliability and heritability in a normative population, we found that these altered network patterns might be due to changes in intrinsic biological factors. Common hubs displayed increased cortical thickness in rBD, decreased cortical thickness in BipM, and no significant differences in cortical thickness in BipD. The spatial distributions of receptors (e.g., NMDA, 5HTT), cellular maps (e.g., L5ET, Micro/PVM, Oligo, Chandelier), and gene expression (PLEKHO1, SCN2A, POU3F2, ANK3) were correlated with these patterns, suggesting potential neurobiological implications for episode dynamics.

Extending previous neuroimaging knowledge in BD, we detailed the functional changes related to episode dynamics, encompassing common and specific patterns. A meta-analysis of twenty-three resting-state functional connectivity studies identified consistent alterations in the DMN, FPN, and SN in rBD (Syan et al. 2018). Our study primarily emphasizes that the limbic network and frontoparietal control network are shared neural representations of bipolar disorder across different stages of the illness. A widely accepted mechanism underlying bipolar disorder is the early developmental disruption in brain networks that regulate emotional behavior (e.g., white matter connectivity and prefrontal pruning), leading to reduced connectivity between the ventral prefrontal network and limbic brain regions, particularly the amygdalaStrakowski et al. (2012). Our research further underscores and confirms the universality of this theory across different stages of the disorder. In addition, we confirmed a specific pattern for rBD in the prefrontal cortex. Other studies comparing functional connectivity between unipolar depression and bipolar depression indicated differences in limbic and prefrontal networks (Anand et al. 2009, He et al. 2016, Liu et al. 2019). Comparisons of episodic and remitted states in BD showed reduced connectivity in the DMN and affective network (Wang et al. 2020). Our comprehensive study provided more detailed insights into episode dynamics in BD, identifying common alterations in the salience-attention network, a specific default mode-salience network pattern for BipM, a sensory-prefrontal network specific to BipD, and a limbic network specific to rBD. These findings suggest that common patterns are less plastic, while specific patterns may be more sensitive to environmental changes. By combining test-retest reliability and heritability metrics in a normative sample, we identified potential neuroimaging markers in the prefrontal cortex as targets for treatment.

Integrating clinical symptoms with specific properties, we summarized the general structure and emphasized the regions in these network patterns. Nodal degree, which describes the connectivity density of brain networks, was related to synaptic plasticity and changes in neuronal connectivity patterns (Tijms et al. 2013). We found that the region degree of the default mode-salience network in BipM positively correlated with manic clinical presentations, leading to instability in decision-making and increased impulsive behavior (Fineberg et al. 2014). Conversely, during depressive episodes, the nodal degree of the DMN and affective network negatively correlated with clinical depressive symptoms, suggesting reduced connectivity density in these brain regions during depression. This aligns with decreased neurotransmitters, such as dopamine (Delva and Stanwood 2021) and glutamate (Bernard et al. 2011), emphasizing the impairment role of these network hubs in depressive disorders. In rBD patients, these findings may indicate neuroplastic changes in limbic and prefrontal networks (Gandhi et al. 2020), potentially correlating with symptom relief. Lastly, the nodal degree of the common salience-attention network negatively correlated with cognitive impairment, highlighting the significant impact of these regions on emotion regulation and cognitive function in BD patients. The consistency between dynamic and static network attribute results confirms the robustness of network lesions over time, supporting the specificity and commonality of different network patterns, especially their hubs in BD across different episode types.

We further examined the structural basis of brain function, which could clarify the long-term effects on common hubs in the VAN across various disease episodes. Structural studies showed a significant decrease in cortical thickness in this region for BipM, a significant increase for rBD, and no difference for BipD, aligning with previous findings (Hanford et al. 2016, Zhu et al. 2022). The functional connectivity results explain the VAN-based compensatory mechanism across different episode types of BD (Chen et al. 2018). The differential structural basis of this functional compensatory change may be related to emotional excitement in BipM, leading to structural damage, i.e., reduced neuronal density, and structural compensation during remission, which may be related to symptom relief and self-regulation mechanisms. The functional differences in BipD did not result in changes in cortical thickness, possibly due to short-term compensatory regulation during depression (Townsend and Altshuler 2012). These results provide valuable insights for subsequent clinical treatments, suggesting that targeted treatments at different episode types may have different side effects.

We also examined the relationship between specific-common networks across various episode types and receptors, finding that all episode-type functional patterns were negatively associated with serotonin receptors and the L5ET map. L5ET cells, associated with executive functions and perceptual information processing (Farsi et al. 2023), and serotonin receptors, involved in central nervous system excitatory behavior and mood fluctuations (Barnes and Sharp 1999), may influence emotion and cognitive imbalance by regulating key nodes within the default mode and limbic networks (Rolls 2015, Smallwood et al. 2021). Dopamine receptors and Oligo maps were also associated with specific-common patterns. Oligo cells are crucial for normal neuronal function(Bechler et al. 2015). D1, D2, DAT, and NET may trigger irregularities in connectivity within the default mode network, limbic network, and ventral attention network across various episode types by regulating dopamine levels and engaging with other neurotransmitter systems, impacting brain network operations related to emotions and cognitive control (Faraone and Radonjić 2023, Kienast and Heinz 2006, Treadway and Pizzagalli 2014). Additionally, NMDA receptors were specifically negatively correlated with rBD, suggesting a role in neuroadaptation and recovery processes (Hansen et al. 2017, Hirschfeld et al. 2007).

Furthermore, BD involves the interaction of multiple genetic factors. Genes such as PLEKHO1, SCN2A, POU3F2, and ANK3 may play significant roles in the genetic basis of the disease (Stahl et al. 2019). These genes may affect emotion regulation and cognitive function by influencing neuron function, synaptic transmission, and neural plasticity pathways (Li et al. 2021), impacting neural function during remission. The connections between manic phase patterns and genes like SCN2A, POU3F2, and ANK3, along with the involvement of PLEKHO1, LMAN2L, SCN2A, and POU3F2 in shared patterns, signify diverse roles across various episodes (Smeland et al. 2020). Shared patterns suggest consistent gene influence over disease progression, while distinct patterns highlight their varying significance across episodes. The absence of specific bipolar genetic expression during depressive phases might imply greater diversity in genetic underpinnings influenced by environmental factors (Saveanu and Nemeroff 2012). These discoveries offer insights into the relationship between genes and phenotypes in BD, aiding in understanding the molecular basis of the disease.

### Limitation

This study has certain limitations that warrant consideration. Firstly, the sample was derived from a single center, and the sample size was relatively small, which may constrain the reliability and generalizability of the results. Future studies should aim to conduct multicenter research with larger sample sizes to address these limitations and enhance the external validity of the findings, thereby increasing their generalizability and applicability to a broader patient population. Secondly, this investigation included both initial BipM patients and those with rBD beyond the medication washout period to gain a comprehensive understanding of the imaging features of BD. The potential impact of medications on brain structure and function is a limitation that must be acknowledged, as it may affect the results. This is a common challenge faced by many studies in this field. Currently, an ideal approach to fully address this issue has not been identified. Lastly, this study employed a longitudinal rating scale to assess symptom changes in BipM patients before and after treatment, while the rest of the study followed a cross-sectional design. Consequently, the study was unable to observe the trajectory of structural and functional neuroimaging changes in the brains of BD patients over time. Future research should consider implementing longitudinal follow-up assessments to capture the variations in brain structure, function, and emotional and cognitive functions over different periods in individuals with BD.

## CONCLUSION

In this study, we utilized structural and functional magnetic resonance imaging data to extract specific and common network patterns associated with different episode types of BD. Unique network patterns were correlated with clinical symptoms specific to each episode type, while common networks were associated with cognition and exhibited different structural bases across various mood episodes. This revealed distinct functional and structural alterations related to the disease. The more robust and heritable patterns were found to be underpinned by different receptors and genes. This study represents the first comprehensive investigation into the mechanisms of bipolar affective disorder across different episode types, encompassing structural, functional, and receptor aspects. Our findings provide a research foundation for developing comprehensive and individualized treatments, ranging from brain network-targeted TMS to gene-based therapies, for individuals with BD.

## METHODS

### Cohort description

This study was approved by the Ethics Committee of Renmin Hospital of Wuhan University (Ethics Approval Number: WDRY22022-K195). Prior to initiating the study, the objectives, procedures, and precautions of the research were thoroughly explained to the participants and their families (due to some patients lacking decision-making capacity). Informed consent was obtained only after ensuring that all participants and their families were fully informed and understood the details. Written informed consent forms were then signed. The study was registered in the National Medical Research Registration Information System and the Chinese Clinical Trial Registry (Registration Number: ChiCTR2200064938).

#### Clinical symptoms and cognitive assessment

The Young Mania Rating Scale (YMRS) (Young et al. 1978) is a clinical assessment tool specifically designed to evaluate manic symptoms and their severity. It consists of 11 items, with scores ranging from 0 to 4 for items 1, 2, 3, 4, 7, 10, and 11, and from 0 to 8 for items 5, 6, 8, and 9. Severity levels of manic symptoms are determined based on the total score: Normal (0-5), Mild (6-12), Moderate (13-19), Severe (20-29), Very Severe (30 and above). Clinicians assess each item based on patient responses, and the total score guides the understanding and management of manic symptoms.

The Hamilton Anxiety Scale (HAMA) (Maier et al. 1988) was initially designed to objectively and reliably assess anxiety symptoms. The revised 1980 version includes 14 items, scored from 0 to 4, representing levels of symptom severity: (0) None, (1) Mild, (2) Moderate, (3) Severe, (4) Very Severe. Anxiety factors are categorized into somatic and psychic, and scores are analyzed accordingly. Chinese criteria categorize scores as (1) Total score ≥29 indicates severe anxiety; (2) ≥21 indicates definite anxiety; (3) ≥14 indicates the presence of anxiety; (4) >7 suggests possible anxiety; (5) <7 indicates an absence of anxiety symptoms.

The Hamilton Depression Rating Scale (HAMD) is widely used in clinical settings for assessing depression severity (Hamilton 1960). The HAMD-17 is a newer version indicating disease severity through lower scores for milder conditions and higher scores for more severe cases. HAMD is categorized into seven factors: (1) Anxiety/Somatization, (2) Weight, (3) Cognitive Disturbances, (4) Diurnal Variation, (5) Retardation, (6) Sleep Disturbances, and (7) Desperation. Chinese criteria categorize scores as: (1) Total score <7 indicates normal; (2) 7-17 suggests possible depressive symptoms; (3) 17-24 indicates definite depressive symptoms; (4) >24 indicates severe depressive symptoms. The Perceived Disability Questionnaire (PDQ) assesses the degree of impairment in daily life, work, social, and leisure activities. It consists of 20 items, each scored on a 5-point scale from 0 (no difficulty) to 4 (extremely difficult). The questionnaire covers various functional activities such as concentration, work completion, social interaction, and household chores.

In this study, two trained doctoral students in psychiatry conducted neurocognitive assessments on all participants in the same environment, with each assessment taking approximately 60 minutes.

#### Subject recruitment

We collected data from patients diagnosed with Bipolar Disorder who sought treatment at the Department of Psychiatry and Psychology, Wuhan University People’s Hospital, from September 2021 to December 2023. Initial screening of research participants was conducted using the Chinese version of the Mini-International Neuropsychiatric Interview (MINI) (Amorim 2000). All participants were diagnosed by two experienced psychiatrists following the Diagnostic and Statistical Manual of Mental Disorders Fifth Edition (DSM-5) criteria for BD (Diagnostic 1994). Inclusion criteria for the BipM group: Age 18-45; DSM-5 diagnosis of BD, YMRS > 7, and HAMD < 7; First episode and untreated or first-time undergoing treatment. The inclusion criteria for the BipD group were HAMD-17 > 7 and YMRS < 7. Inclusion criteria for the rBD group: Age 18-45; DSM-5 diagnosis of BD, HAMD < 7, and YMRS < 12; Patient-initiated discontinuation for more than 14 days. Exclusion criteria: MRI contraindications; Organic brain diseases; Other mental illnesses; History of medication or physical therapy for BipM patients; Left-handedness; Unstable physical illnesses; Substance abuse history; Pregnancy or lactation; Concurrent other brain function disorders.

HCs were recruited through community, university, and Hubei Provincial People’s Hospital posters, with age and sex to the patient group, and right-handedness. Exclusion criteria for HCs: MRI contraindications; Organic brain diseases; Substance abuse history; Pregnancy or lactation; Family history of neurological or psychiatric disorders. All participants had withdrawal and termination criteria: Withdrawal criteria included voluntary withdrawal of informed consent and researcher judgment of unsuitability to continue. Termination criteria involved non-cooperation leading to data invalidation, exclusion from final data analysis, and discontinuation of further investigation. Noted that there were not significant difference among rBD, BipD, BipM and HC in age, sex and handness (p > 0.05). But there were significant difference in HAMD (F = 321.05, p = 1.46e-47), YMRS (F = 111.61, p = 1.42e-27), PDQ (F = 15.67, p = 9.75e-7) and HAMA (F = 35.67, p = 9.32e-13).

#### Imaging

The research participants’ collection procedures and data quality control can be seen in the Supplementary material (Figure.S1). MRI images were acquired using an Achieva 3T MRI scanner (GE, SIGNA Architect) equipped with a 48-channel head coil. Participants were instructed to stay awake, remain motionless, relax, and keep their eyes closed during the scanning procedure. To minimize head movement and reduce scan noise, foam padding, and soft earplugs were provided. All scans were performed by two licensed MRI technicians with intermediate professional titles. T1_3D data sets were acquired with a maximum set repetition time, minimum set echo time, a NEX of 1, a layer thickness of 2mm, and a field of view of 256 × 256 mm^2^. And scan time was 7 minutes. For rs-fMRI, a TR of 2000ms, a TE of 30ms, a FOV of 220mm × 220mm, a flip angle of 90°, a matrix of 64 × 64, a resolution of 3 × 3 × 3, a slice thickness of 36, and 240-time points were acquired.

### Data preprocessing and Functional connectome

For all datasets, raw DICOM files were converted to Brain Imaging Data Structure (BIDS) format (Gorgolewski et al. 2016) using HeuDiConv v0.13.1 (Esteban et al. 2019). The structural and functional preprocessing of all image data were performed with fMRIPrep 23.0.2 (Esteban et al. 2019), which is based on Nipype 1.8.6 (Gorgolewski et al. 2011). The main anatomical data preprocessing steps include intensity normalization, brain extraction, tissue segmentation, surface reconstruction, and spatial normalization (Dong et al. 2018). The main functional data preprocessing steps include head motion correction, slice-time correction, and coregistration. For original preprocessing details generated by fMRIPrep (see (Esteban et al. 2019)). The derived functional time series were parcellated into the Schaefer200×7 atlas (Schaefer et al. 2018) and underwent a confound removal process implemented in Nilearn via adopting the “simple” strategy from (Wang et al. 2024). The whole process includes high-pass filtering, motion and tissue signals removal, detrending, and z-scoring. Functional connectivity matrices were then estimated for each subject using the zero-lag Pearson correlation coefficient.

### Common network pattern and specific network pattern

To identify network changes, we first compared the functional connectome between BipM, BipD, and rBD patients with healthy controls using two-sample t-tests with False Discovery Rate (FDR, threshold = 0.05), controlling for age and sex as covariates. We delineated common and specific network patterns across the disease cohorts by identifying overlapping and non-overlapping sets of edges within differential network profiles. Notably, specific disease network patterns could diagnose particular episode types better compared with rest two types, whereas common patterns could identify all three episode types. To validate the specificity of these network patterns, we employed machine learning techniques to assess their capacity to discern variations across different disease cohorts. Specifically, we used a Support Vector Machine (SVM) with a radial basis kernel and default parameters to classify patients and healthy controls. The data were split into training and testing sets using a leave-one-out approach, and network patterns were selected based on specific and common network profiles. Model performance was evaluated using accuracy and the Area Under the Curve (AUC). Additionally, we built a classification null model for comparison by randomly selecting the same number of edges consistent with the specific and common networks in disease and normal individuals to compose network patterns. We used SVM classification to obtain accuracy and repeated the entire process 1000 times.

### Reliability and heritability of functional connectivity

Next, we estimated the stability of functional connectivity from the two aspects including test-retest reliability and heritability, which reflect intra-subject robustness across time and biological robustness determined by behavioral genetics. To do so, we utilized the Human Connectome Project (HCP) S1200 data release for the large sample size of a healthy adult population (Van Essen et al. 2013). This dataset comprises four sessions of resting-state functional magnetic resonance imaging (rs-fMRI) scans from 1206 healthy young adults, along with pedigree information, including 298 monozygotic and 188 dizygotic twins, as well as 720 singletons. We specifically selected individuals with a complete set of four fMRI scans that met the HCP quality assessment criteria (Glasser et al. 2013, Van Essen et al. 2012). Ultimately, our sample encompassed 1014 subjects (470 males) with an average age of 28.7 years (range: 22–37). We assessed the test-retest reliability of FC by calculating the intra-class correlation coefficient (ICC) (Shrout and Fleiss 1979). The ICC is defined as 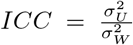, where 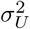 denotes the intra-subject variance and 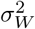 the total variance. Total variance is the sum of intra-subject variance and inter-subject variance. To interpret the results, we calculated the ratio of significant edges among total edges after fdr correction.

To map the heritability of FC in humans, we used the Sequential Oligogenic Linkage Analysis Routines (SOLAR, v8.5.1b)(Almasy and Blangero 1998). In brief, heritability indicates the impact of genetic relatedness on a phenotype of interest. SOLAR uses maximum likelihood variance decomposition methods to determine the relative importance of familial and environmental influences on a phenotype by modeling the covariance among family members as a function of genetic proximity (Almasy and Blangero 1998, Wan et al. 2022). Heritability (i.e. narrow-sense heritability *h*^2^) (Almasy and Blangero 1998) represents the proportion of the phenotypic variance 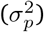 accounted for by the total additive genetic variance 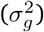, that is 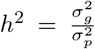. Phenotypes exhibiting stronger covariances between genetically more similar individuals than between genetically less similar individuals have higher heritability. In this study, we quantified the heritability of functional edge from common and specific network patterns. We added covariates to our models including age, sex, age^2^, and age×sex. We calculated heritability as well as the ratio of significant edges among total edges after fdr correction in all two sessions and averaged them as final heritability.

Differential indicators of seed-based functional connectivity (sFC) can establish a potential relationship among specific regions and other rest of brain regions (Wu et al. 2018). Additionally, fMRI is known to be susceptible to environmental noise (Liu 2016) and exhibits substantial individual variability (Dubois and Adolphs 2016). Therefore, considering the noise and instability of functional connections, we compared the robustness and heritability of different regions in the disease compared to healthy adults in HCP. We filtered to identify a few edges with high stability and heritability (p < 0.05). Subsequently, the number of filtered edges that each node possesses was used to define hubs. Hubs with the top 5 filtered edge counts were selected as seeds to construct the network connection model with other brain regions. A representative set of specific and common cortical patterns was then averaged.

### Predict clinical symptom via precise specific network patterns

To further link the extracted specific networks to clinical symptoms and validate the robust biological significance of specific episode patterns, we predicted various clinical measurements using specific patterns based on a multivariate model. Specifically, we trained a support vector regression (SVR) model to predict YMRS, HAMD, and HAMA scores using the specific networks of BipM, BipD, and rBD(Liu et al. 2021). To enhance the variability of clinical information, we combined YMRS, HAMD, and HAMA scores from all 117 samples, including those with BipM, BipD, and rBD. We divided all specific functional edges into training and testing datasets via 10-fold cross-validation to avoid model overfitting. For each iteration, we trained the SVR model (https://www.csie.ntu.edu.tw/~cjlin/libsvm/) with default parameters and a linear kernel. We combined the predicted labels from each fold and estimated prediction accuracy by calculating Pearson’s correlation between predicted and actual labels. To test the robustness of the model, we repeated the entire process 1000 times and averaged all predicted labels from each iteration to obtain the final results.

### Static and dynamic properties of network patterns

Node degree is a crucial metric indicating the extent of a node’s connections within a network, thereby serving as a measure of node centrality and reflecting its significance in network topology. A comprehensive examination of node degree provides insights into the regional functionality within diseased brain networks. Initially, we extracted the top 10% of the connectivity matrix, binarized it, and computed node degrees using the Brain Connectome Toolbox (BCT, https://sites.google.com/site/bctnet/) (Rubinov and Sporns 2010).

The pathology of mental disorders can be conceptualized as the dynamic switching of multiple resting networks (Menon 2011). We employed a sliding window method to segment the BOLD time series into short segments (Hindriks et al. 2016). Subsequently, we calculated the whole brain FC matrix for each segment. Using a time window of 50 TRs in length and a step of 1 TR, the data were segmented. At each window, we computed the Pearson correlation coefficient between the BOLD time courses for each pair of Regions of Interest (ROIs), resulting in a dynamic functional connectome (dFC). The dFC matrix obtained was Fisher’s z-transformed correlation coefficient matrix of all time points, with dimensions of 200 × 200 × 182, where 200 represents the number of ROIs and 182 indicates the number of time windows.

Subsequently, we estimated the dynamic network switch rate to summarize the transition of each ROI across time points (Pedersen et al. 2018). The switch rate, determined using an iterative and ordinal Louvain algorithm, represents the percentage of time windows during which brain nodes transition among various network assignments (Pedersen et al. 2018).

### The clinical characterization of static and dynamic network properties

To investigate the clinical manifestations associated with the extracted networks from different patients, we proceeded to establish a connection between dynamic and static properties and clinical questionnaires. Initially, we identified regions with the highest number of significant edges as hubs and extracted their network properties. Subsequently, we computed Pearson’s correlation between the switch rate and the degree of these hubs in BipM, BipD, rBD, and their common regions with respect to YMRS, HAMA, HAMD, and PDQ, respectively.

### Structural foundation of functional hubs

To explore whether functional hubs exhibit similar structural foundations, we extracted cortical thickness data corresponding to these hubs for each individual. We posited that the pattern shared by the three diseases should demonstrate stability compared to specific hubs and therefore compared these with the common hubs. Subsequently, we computed the average cortical thickness values across these shared hubs within network patterns for each patient, simplifying statistical analyses. This approach facilitated an assessment of the overarching structural mechanisms governing these functional hubs. Following this, we conducted two-sample t-tests to compare the average cortical thickness among individuals with rBD, BipD, and BipM, with age and sex as covariates.

### Receptor maps from Positron Emission Tomography

Receptor densities were assessed through PET tracer investigations covering a total of 18 receptors and transporters across nine neurotransmitter systems. These data were recently shared by Hansen and colleagues (Hansen et al. 2022, Markello et al. 2022)(https://github.com/netneurolab/hansen_receptors). The neurotransmitter systems include dopamine (D1, D2, DAT), norepinephrine (NET), serotonin (5-HT1A, 5-HT1B, 5-HT2, 5-HT4, 5-HT6, 5-HTT), acetylcholine (*α*4*β*_2_, M1, VAChT), glutamate (mGluR5), GABA (GABAA), histamine (H3), cannabinoid (CB1), and opioid (MOR). Volumetric PET images were aligned with the MNI-ICBM 152 nonlinear 2009 (version c, asymmetric) template. These images, averaged across participants within each study, were subsequently parcellated into the Schaefer 200 template. We combined and weighted the averaged receptors/transporters exhibiting more than one mean image of the same tracer (e.g., 5-HT1B, D_2_, VAChT).

### Cellular Maps

Here, we further correlated patterns observed in three episodes of bipolar disorder with 24 cellular maps from a previous study (Zhang et al. 2023). The molecular signature profiles of all cell classes were constructed from snDrop-seq samples provided by Jorstad et al. (2023). Then cell type fractions were deconvolved from microarray samples downloaded from Allen Human Brain Atlas (AHBA; http://human.brain-map.org/) (Shen et al. 2012). The 24 cell types are Lamp5, Pax6, Vip, Sncg, Lamp5, Lhx6, L5ET, L5/L6 NP, L6 CT, L6b, Astro, VLMC, Endo, Micro/PVM, Oligo, OPC, L2/3 lT, L6 lT Car3, L4 lT, L6 lT, L5 lT, Chandelier, Pvalb, Sst, and Sst Chodl, as detailed in Jorstad et al. (2023), Zhang et al. (2023).

### Genetic expression mechanisms

In order to study how the network patterns are regulated by genes, we combined AHBA and the precise brain connectivity pattern for analysis. Regional microarray expression data were obtained from six post-mortem brains. We used the *abagen* toolbox(https://github.com/netneurolab/abagen) (Markello et al. 2021) to process and map the data to 200 parcellated brain regions from Schaefer parcellation. We first extracted the gene expression (Stahl et al. 2019) associated with risking of bipolar disorder from ENIGMA toolbox (Larivière et al. 2021), and finally calculated the correlation between the brain maps of these gene expression and the specific and common connection patterns of different episode types of bipolar disorder.

### Null model

In our current investigation, a persistent inquiry revolves around the topographic correlation existing between common-specific network patterns and other salient features. To delineate these correlations, we employed a null model meticulously designed to systematically disrupt the relationship between two topographic maps while preserving their spatial autocorrelation. Initially, we shuffled parcellation locations to randomize receptor maps according to the methodology outlined in Hansen et al.(Hansen et al. 2022), subsequently calculating the relationship between this map and common-specific network patterns. These resulting spatial coordinates formed the basis for generating null models through the application of randomly-sampled rotations and the reassignment of node values based on the nearest resulting parcel, a process iterated 1000 times. Notably, the rotation was initially applied to one hemisphere and then mirrored onto the other hemisphere. It is noteworthy that the 95th percentile of shuffling occurrence frequencies derived from spatial null models was designated as the threshold value.

## Data Availability

All data produced in the present study are available upon reasonable request to the authors

## DATA AVAILABILITY

The clinical data could be accessed through reasonable requests made to the corresponding authors. The raw fMRI data and MRI data for HCP are available on https://db.humanconnectome.org/. Heritability analyses were performed using Solar Eclipse 8.5.1b (https://www.solar-eclipse-genetics.org). *neuromaps* is available on (https://netneurolab.github.io/neuromaps/usage.html), ENIGMA toolbox is available on (https://enigma-toolbox.readthedocs.io/en/latest/pages.html).

## CODE AVAILABILITY

Code will be available on (https://github.com/Laoma29/Publication_codes).

## ACKNOWLEDGMENTS

Xiaobo Liu is supported by China Scholarship Council. Bin Wan is supported by International Max Planck Research School on Neuroscience of Communication: Function, Structure, and Plasticity (IMPRS NeuroCom), Graduate Academy Leipzig, and Mitacs Globalink Research Award. ZQL acknowledges support from the Fonds de Recherche du Québec – Nature et Technologies (FRQNT).This work was supported by in part by the Health of Hubei Province Scientific Research Project under Grant 2020Cfb512, and project of Mental Health Research Institute of Three Gorges University: YCXL-23-11.

## Supplemental Materials

