## Supplemental Materials for "Episode-specific and common intrinsic functional network patterns in bipolar"

1.Department of Psychiatry, Wuhan Wuchang Hospital, Wuhan University of Science and Technology, Wuhan 430063, China
2.McConnell Brain Imaging Centre, Montr\'eal Neurological Institute, McGill University, Montreal, Canada
3.Max Planck Institute for Human Cognitive and Brain Sciences, Leipzig, Germany
4.Institute of Neuroscience and Medicine (INM-7: Brain and Behavior), Research Center Jülich, Jülich, Germany
5.Department of Psychology, Yale University, New Haven, CT, USA
6.Department of Human Genetics, McGill University, Montreal, Canada
7.Sichuan Provincial Center for Mental Health, Sichuan Provincial People's Hospital, School of Life Science and Technology, University of Electronic Science and Technology of China, Chengdu, China
8.First Hospital of Quanzhou Affiliated to Fujian Medical University, Quanzhou, Fujian, China
9.Peking University Sixth Hospital, Peking University Institute of Mental Health, NHC Key Laboratory of Mental Health, Beijing 100191, China
10.Department of Neurology, Renmin Hospital of Wuhan University, Wuhan, China
+ Equal contribution
*Corresponding author

### Diagnostic criteria

### The research participants collection procedures and data quality control

The study excluded 92 participants for various reasons. In the BipD group, exclusions included 9 participants who were ultimately diagnosed with major depressive disorder during follow-up, 1 with brain organic diseases, 2 unable to persist in scanning, 4 with excessive head movement, and 4 with metal braces. The BipM group saw the exclusion of 8 participants due to excessive head movement, 2 who were misdiagnosed during follow-up, 4 with comorbid sex identity disorder, 2 with brain organic diseases, and 8 unable to persist in scanning. In the rBD group, 2 participants were excluded due to a follow-up diagnosis of major depressive disorder. In the HCs group, exclusions were 4 participants with metal braces, 4 with head motion exceeding 2mm in any direction who did not consent to rescanning, and 15 with a significant family history of psychiatric or neurological disorders among first-degree relatives. Ultimately, all 38 BipM patients completed follow-up through various methods. The final analysis included 42 BipD patients, 38 BipM patients, 37 rBD patients, and 35 HCs (Fig. S1).

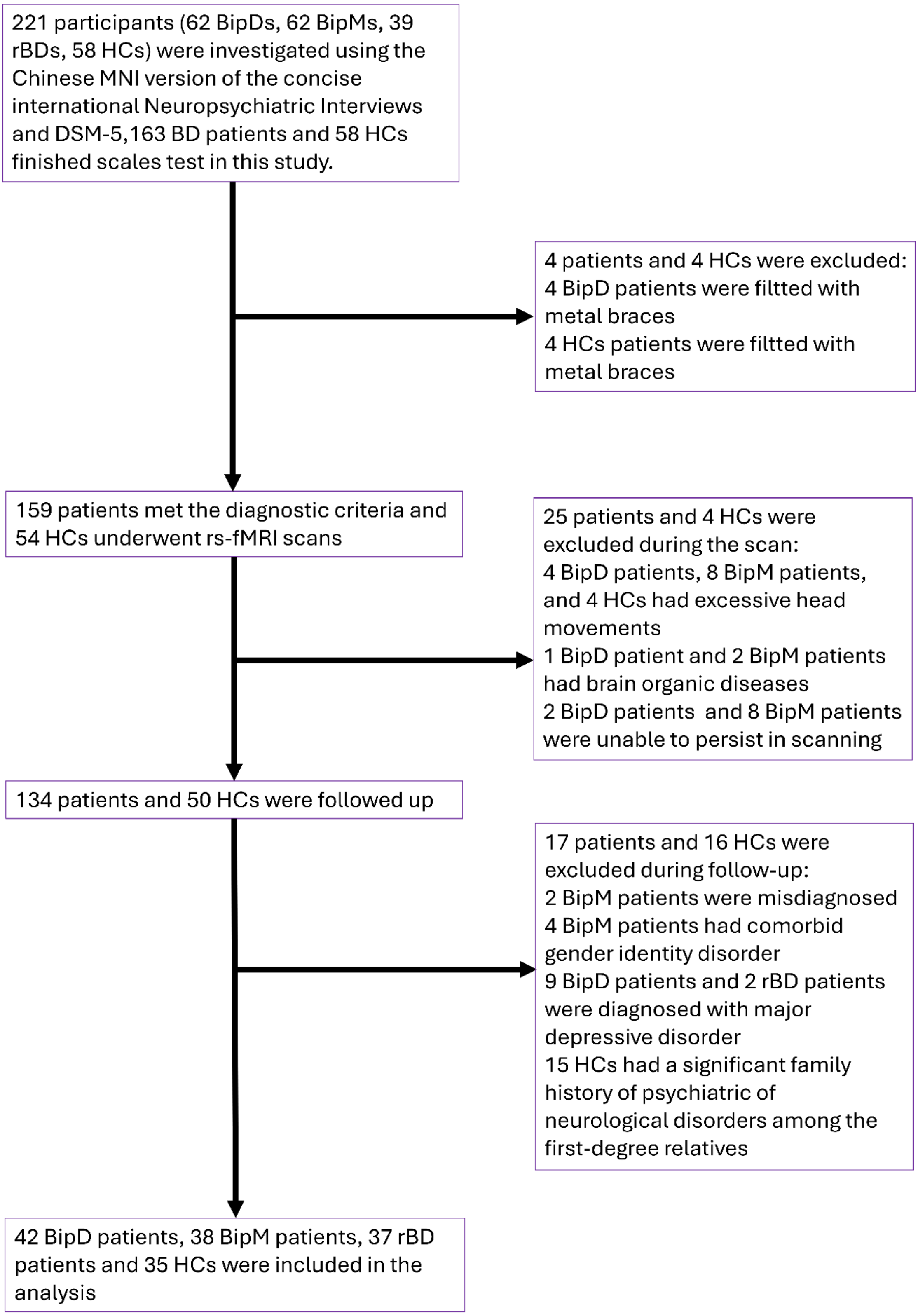


S-Figure.1. The research participants collection procedures and data quality control

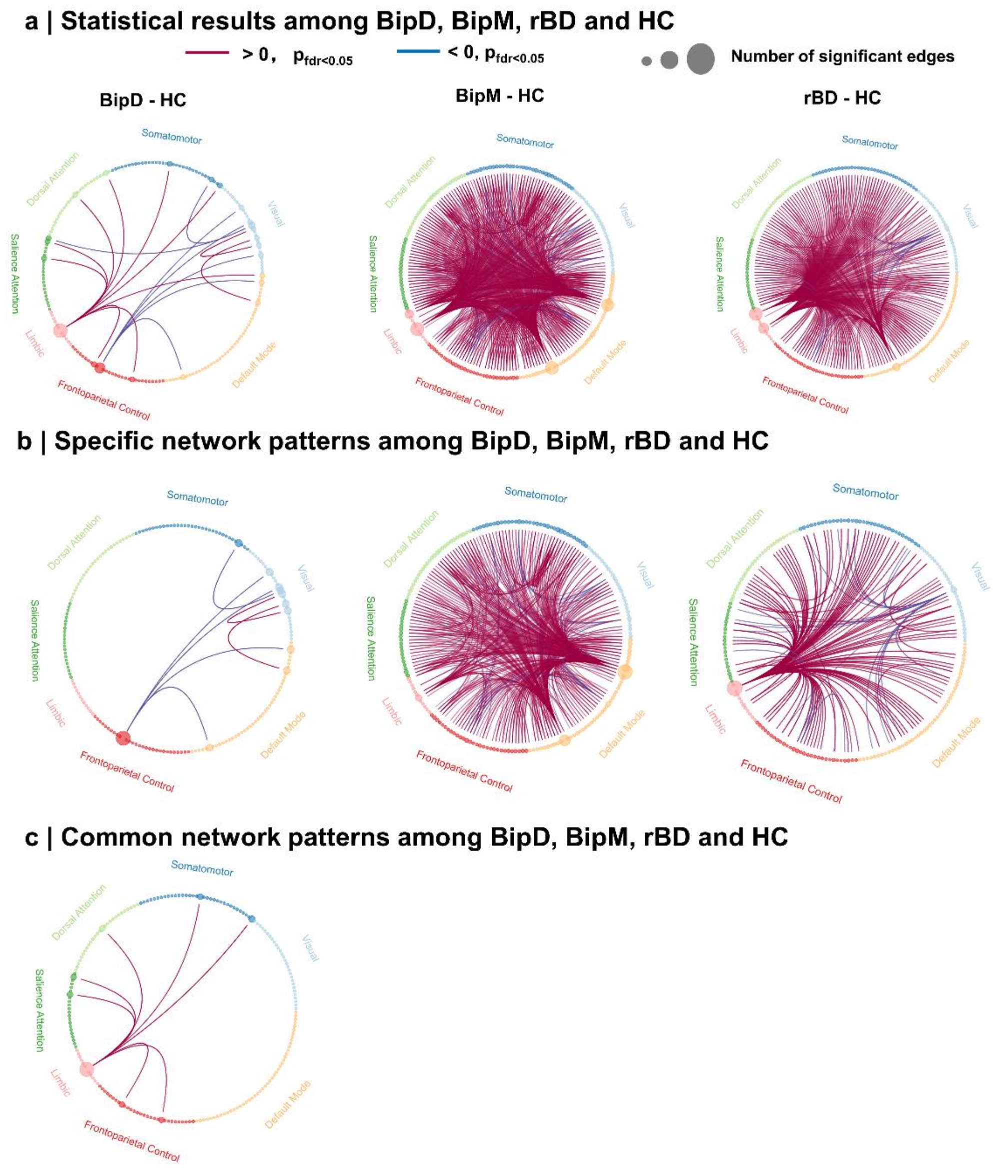


S-Figure.2.The significant different disease and healthy control.

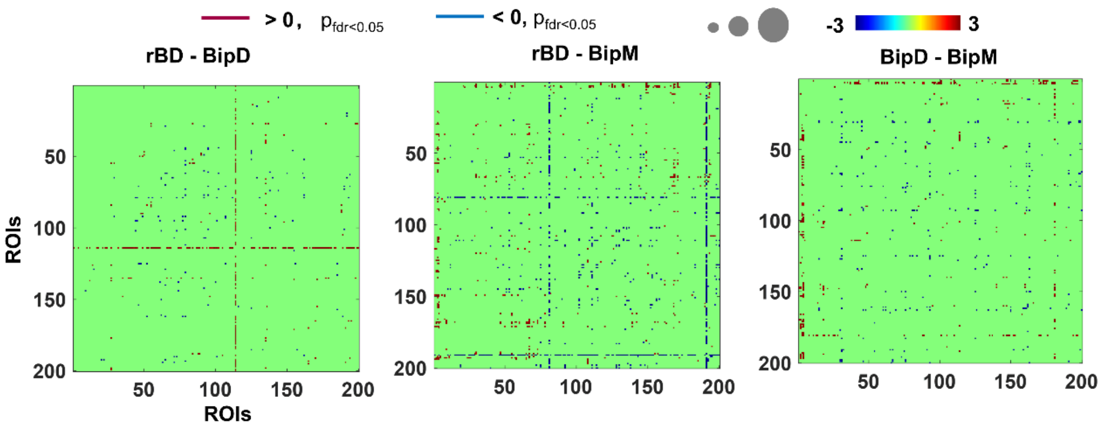


S-Figure.3.The significant comparison connectome matrix among different phase of bipolar disorder.

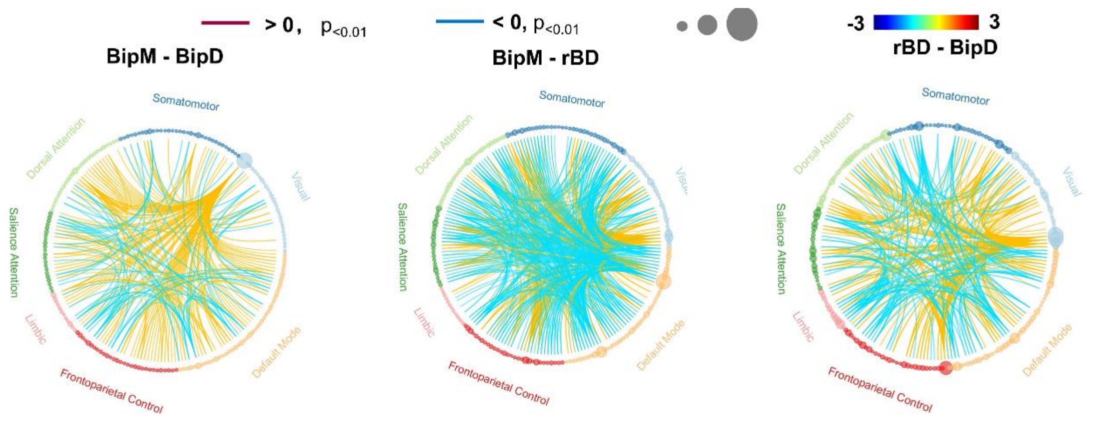


S-Figure.4.The significant comparison network patterns among different diseases.

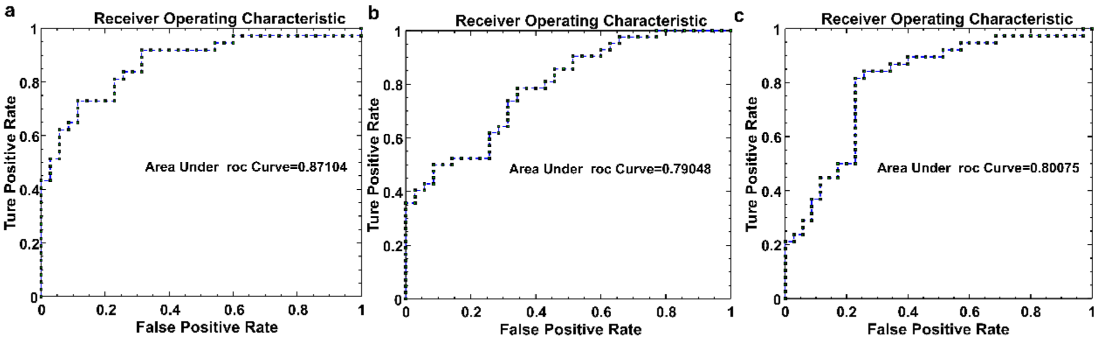

S-Figure.5.ROC Curve for classification between BipD, BipM, rBD and HC.

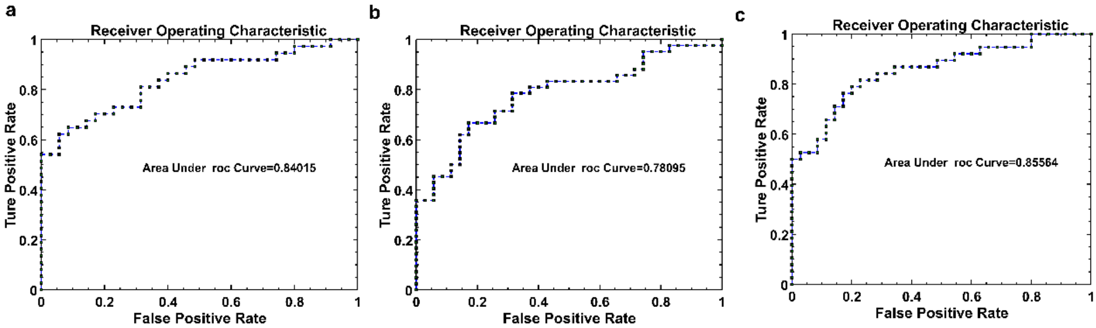


S-Figure.6.ROC Curve for classification between BipD, BipM, rBD and HC.

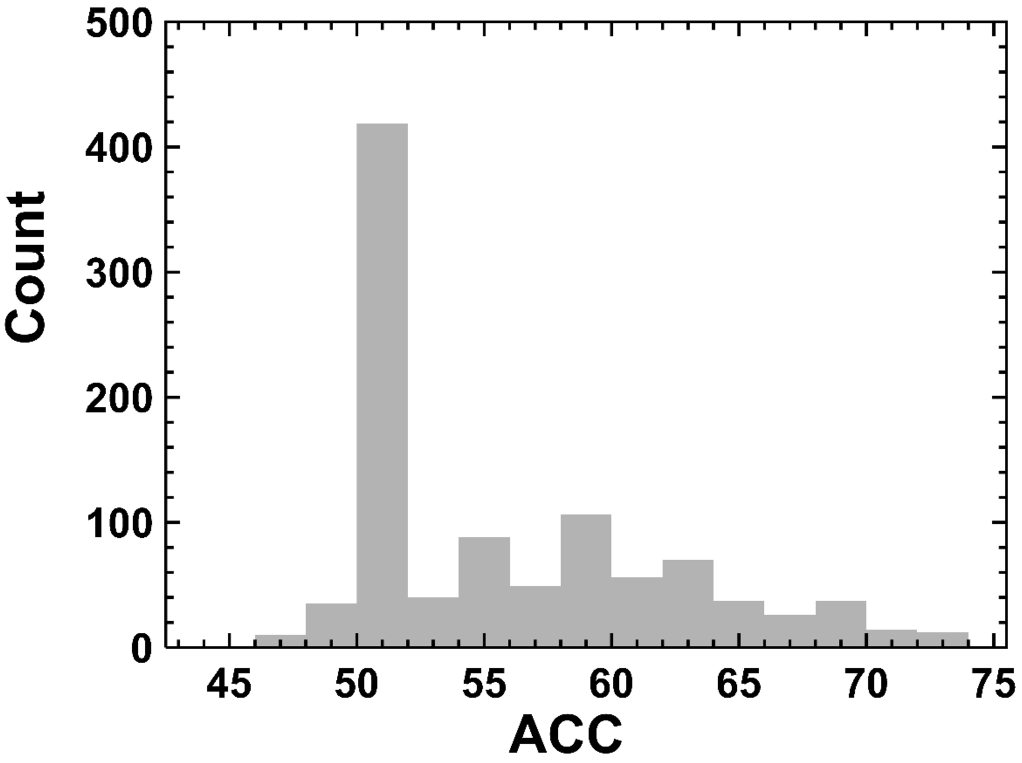


S-Figure.7. null model for classification.

Table. S-1 The classification between specific network patterns of patients and healthy control

| | Disease stage | Accuracy (%) | Sensitivity (%) | Specificity (%) | AUC |
|---|---|---|---|---|---|
| BipD-Specific | BipD - HC | 80.56 | 91.89 | 69.57 | 0.8710 |
| | BipM - HC | 59.74 | 66.67 | 51.43 | 0.6884 |
| | rBD - HC | 56.16 | 57.89 | 54.29 | 0.6060 |
| BipM-Specific | BipD - HC | 51.39 | 100 | 0 | 0.6224 |
| | BipM - HC | 71.43 | 85.71 | 54.29 | 0.7905 |
| | rBD - HC | 52.05 | 100 | 0 | 0.6399 |
| rBD -Specific | BipD - HC | 51.39 | 100 | 0 | 0.50 |
| | BipM - HC | 58.44 | 73.81 | 40 | 0.6354 |
| | rBD - HC | 75.34 | 89.47 | 60 | 0.8008 |

Table. S-2 The classification between shared network patterns of patients and healthy control

| | Disease stage | Accuracy (%) | Sensitivity (%) | Specificity (%) | AUC |
|---|---|---|---|---|---|
| Shared | BipD - HC | 75 | 70.27 | 77.14 | 0.8401 |
| | BipM - HC | 72.73 | 76.19 | 68.57 | 0.7810 |
| | rBD - HC | 79.45 | 76.32 | 82.86 | 0.8556 |

Table. S-3 Nodes for specific network pattern of BiPM

| Regions | *X* | *Y* | *Z* | *Number of significant edges* | *P<0.05* |
| --- | --- | --- | --- | --- | --- |
| RH_Default_PFCdPFCm_1 | 208 | 62 | 76 | 147 | |
| LH_Default_PFC_2 | 205 | 63 | 74 | 84 | |
| RH_Default_PFCdPFCm_2 | 208 | 62 | 77 | 39 | |
| RH_Limbic_OFC_1 | 224 | 249 | 165 | 24 | |
| LH_Default_PHC_1 | 206 | 62 | 79 | 16 | |

Table. S-4 Nodes for specific network pattern of BiPD

| Regions | *X* | *Y* | *Z* | *Number of significant edges* $P<0.05$ |
|---|---|---|---|---|
| LH_Cont_pCun_1 | 231 | 149 | 35 | 4 |
| LH_Vis_9 | 120 | 18 | 138 | 1 |
| LH_Vis_11 | 120 | 18 | 140 | 1 |
| LH_Vis_14 | 120 | 19 | 135 | 1 |
| LH_SomMot_4 | 70 | 130 | 177 | 1 |

Table. S-5 Nodes for specific network pattern of rBD

| Regions | *X* | *Y* | *Z* | *Number of significant edges* | *P<0.05* |
|---|---|---|---|---|---|
| LH_Limbic_OFC_2 | 220 | 248 | 166 | 93 | |
| RH_Vis_1 | 124 | 18 | 129 | 16 | |
| LH_Vis_11 | 120 | 18 | 140 | 4 | |
| RH_Cont_Par_1 | 234 | 147 | 35 | 4 | |
| LH_Cont_Cing_2 | 231 | 149 | 37 | 3 | |

Table. S-6 Nodes for shared network patterns of BiPM, BiPD and rBD

| Regions | *X* | *Y* | *Z* | *Number of significant edges* | *P<0.05* |
|---|---|---|---|---|---|
| RH_Limbic_OFC_1 | 224 | 249 | 165 | 7 | |
| LH_SomMot_1' | 70 | 130 | 174 | 1 | |
| LH_DorsAttn_PrCv_1 | 0 | 119 | 16 | 1 | |
| LH_SalVentAttn_ParOper_2 | 196 | 58 | 252 | 1 | |
| LH_SalVentAttn_FrOperIns_4 | 196 | 59 | 254 | 1 | |
